# Bidirectional associations between cannabis use, oddball performance and P3 event-related potential

**DOI:** 10.64898/2026.06.09.26355188

**Authors:** Christina E. Garasky, Kellyn Spychala, Fanghong Dong, Andrey P. Anokhin, Ryan Bogdan, Grace Chan, Victor Hesselbrock, Chella Kamarajan, Sivan Kinreich, Sally I-Chun Kuo, Jodi Kutzner, Alex P. Miller, Ashwini K. Pandey, Gayathri Pandey, Martin H. Plawecki, Jessica E. Salvatore, Marc Schuckit, COGA Collaborators, Kathleen K. Bucholz, Vivia V. McCutcheon, Bernice Porjesz, Jacquelyn Meyers, Arpana Agrawal

## Abstract

**Importance:** Cannabis use remains prevalent in youth despite concerns regarding its potential impact on cognitive function. Unraveling whether the association between cannabis use and cognition is partially due to preexisting differences or primarily related to use is vital to understanding underlying mechanisms.

**Objective:** To estimate the longitudinal association between cannabis initiation and cognitive trajectories, indexed by task performance and P3 event-related potential (ERP), and to estimate whether baseline cognition is associated with cannabis initiation.

**Design:** Data were analyzed from the ongoing longitudinal Collaborative Study on the Genetics of Alcoholism (COGA) cohort, which was followed up approximately every 2-5 years from 2004 to 2025.

**Setting:** 6 sites across the United States.

**Participants:** Adolescent and young adult offspring of past COGA participants and control families who reported on their cannabis use and who had Visual Oddball (VOP) performance and P3 ERP data (N=4814; 52.4% female, 68.4% white) were grouped based on the timing of cognitive data collection relative to cannabis initiation into Pre-onset (n=2,449; ≥1 assessment) and Post-onset (n=998; ≥3 assessments) subsamples.

**Main Outcomes and Measures:** VOP measures include performance accuracy (%), reaction times (ms), and P3 amplitude (μV) and latency (ms) during target trials. Cannabis measures included lifetime use of cannabis (i.e., ever used) and age at first use.

**Results:** High P3 amplitude, and prolonged P3 latency and reaction time were associated with a reduced hazard of cannabis initiation (All Hazards Ratio, [H.R.s]< 0.91, p’s<.008). Following initiation, cannabis use was associated with steeper declines in P3 amplitude (b=-0.29, p=0.02) and stabilized reaction time (b=0.35; p=0.005). Steeper decline in P3 amplitude (i.e., slope) was associated with greater cannabis progression (e.g., Cannabis Use Disorder, Odds Ratio, [O.R.]=2.34, p<.001), whereas steeper decline in reaction time was associated with reduced progression (O.R.=.79, p=.002).

**Conclusion:** Baseline P3 indices and reaction time were associated with cannabis initiation, while cannabis use was associated with subsequent changes in P3 amplitude and reaction time trajectories. These findings indicate that accelerated neurodevelopment may modify the likelihood of cannabis initiation which, in turn, may further contribute to neurocognitive changes that deepen cannabis involvement.

**Key Points:** *Question:* Are associations between cannabis use and cognition attributable to preexisting neurodevelopmental differences, related to cannabis use, or reciprocal processes?

*Findings:* In this longitudinal cohort study of participants from the Collaborative Study on the Genetics of Alcoholism (COGA), high P3 amplitude and prolonged P3 latency and reaction time were associated with reduced risk of cannabis initiation. Following initiation, cannabis use was associated with steeper declines in P3 amplitude and faster but stabilized reaction time.

*Meaning:* Accelerated neurodevelopment may modify the likelihood of cannabis initiation which, in turn, further contribute to neurocognitive changes that deepen cannabis involvement and make it problematic.

---

Cannabis is among the most commonly initiated substances during adolescence with only 14% of 12th graders associating any risk with experimentation.^1,2^ Cannabis use, particularly when prolonged, symptomatic, and early-onset, is associated with adverse behavioral outcomes including impaired cognition and broad spectrum psychiatric risk.^3–9^ Cannabis use also alters neural activuty,^10–15^ with some effects appearing dose dependent.^11,13,16–20^ For example, delta-9-tetrahydrocannobinol (THC) exerts effects on the endocannabinoid system, a neuromodulatory system involved in neural communication and refinement,^21–23^ and alters dopaminergic, GABA, and glutamate activity critical for healthy neurodevelopment.^24,25^ In conjunction, the peak period of cannabis initiation is youth,^26^ a period of rapid brain maturation during which cognitive processes are developing^27^. Thus, prevailing neurobiological models implicate both “top-down” corticolimbic cognitive processes and “bottom-up” prefrontal affective processes in adolescent substance use.^28^ What is less well-understood is whether preexisting cognitive differences are associated with initiating cannabis use, thus establishing a cycle of onset and escalating use that further deepens cognitive decline.^15^ Despite bidirectional hypotheses regarding cannabis use and cognition, the vast majority of prior research has been cross-sectional, with few well-powered longitudinal studies able to assess neural activity before and after cannabis onset. Clarifying whether neurocognitive differences (e.g., P3 indices) precede cannabis initiation, are related to canabis use, or reflect a reciprocal process may provide important insight into the mechanisms underlying cannabis involvement and its developmental course.^29^

Longitudinal studies enriched for cannabis onset are a valuable resource for disentangling preexisting cognitive differences from cognitive changes associated with cannabis use.^16,30–33^ While there are many approaches to measuring cognition, electroencephalogram (EEG) derived neurophysiological indices like the P3 event-related potential (ERP) observed during tasks like the Visual Oddball,^34^ offer a non-invasive, relatively inexpensive, reliable, and temporally specific approach to examining the neurophysiology of substance use.^35–37^ The P3 (or P300) is a positive-going ERP elicited by rare oddball (target) stimuli that require a speeded response and to ignore intermixed, more frequent, and standard (non-target) stimuli. The oddball P3 peaks 300-600 ms post-stimulus with maximum amplitude in the mid-parietal area of the scalp^38^, and is believed to reflect updating working memory and internal representation of the task context and allocation of attention to the target stimulus.^39–41^ Consistent with these definitions, greater amplitudes and shorter latencies are generally associated with superior cognition.^42–45^

Acute cannabis intoxication is associated with decreases in P3 amplitude and increases in P3 latency.^46,47^ Individuals who use cannabis also display similarly low P3 amplitude and prolonged latency relative to those who do not use cannabis.^16,17^ While P3 activity is strongly associated with cognitive task performance (e.g., reaction, accuracy, error), cannabis has been shown to alter neural activity even in the absence of behavioral change^48^ demonstrating the utility of neurophysiological biomarkers associated with cannabis use.^49^ Related research has also shown that preexisting lower P3 amplitude is associated with the subsequent onset of alcohol use.^50,51^ Although no studies to date have examined the impact of preexisting P3 variability in cannabis initiation, it is plausible that P3-indexed cognition serves as both a marker for the cannabis initiation and is also modified by consequent substance use.

To address these gaps, the current study aimed to utilize longitudinal cannabis use, Visual Oddball Paradigm (VOP) P3 and task performance data from the Collaborative Study on the Genetics of Alcoholism (COGA), to differentiate preexisting and cannabis-related cognitive differences that are associated with use, and assess whether cannabis-related cognitive changes were more likely to be associated with cannabis use progression. We hypothesized that lower baseline amplitudes and accuracy, and prolonged latencies and reaction times would be associated with increased hazards of cannabis initiation. Following cannabis use, we hypothesized that steeper declines in P3 amplitudes and accuracy, and prolonged latencies would be associated with use of cannabis, but that response times would be unlikely to change due to habituation effects of multiple assessments.

## Method

### Sample

Data were drawn from the longitudinal component of the COGA^52–54^ (see Supplemental Note for details). Briefly, offspring of the original COGA cohort were recruited at ages 12-22 years and reassessed every 2 years, with additional participants added during a later-life follow-up study (∼5 years later). Data included individuals who (1) completed cannabis use questions in the adolescent or adult Semi-Structured Assessment for the Genetics of Alcoholism (SSAGA),^55,56^ and (2) had performance and P3 ERP data (N=4814;**Figure 1**). To distinguish cognitive differences before versus after cannabis initiation, participants were divided into “Pre-onset” (N=2449) and “Post-onset” (N_3+_=998;N_4+_=585) analytical subsets. The Pre-onset subset included participants with ≥1 P3 assessment (averaged if >1) and was used to assess cognition prior to initiation through survival modeling. The Post-onset subset included participants with ≥3 P3 assessments and was used to assess post-initiation cognitive change through latent growth modeling. For participants reporting lifetime cannabis use, only P3 assessments occurring before (for Pre-onset subset) or after (for Post-onset) cannabis initiation were included. Post-hoc analyses, assessing within-person change associated with cannabis use, included participants with ≥3 P3 assessments regardless of cannabis initiation timing (N=1119). Representative analyses were conducted across the samples.

**Figure 1.**
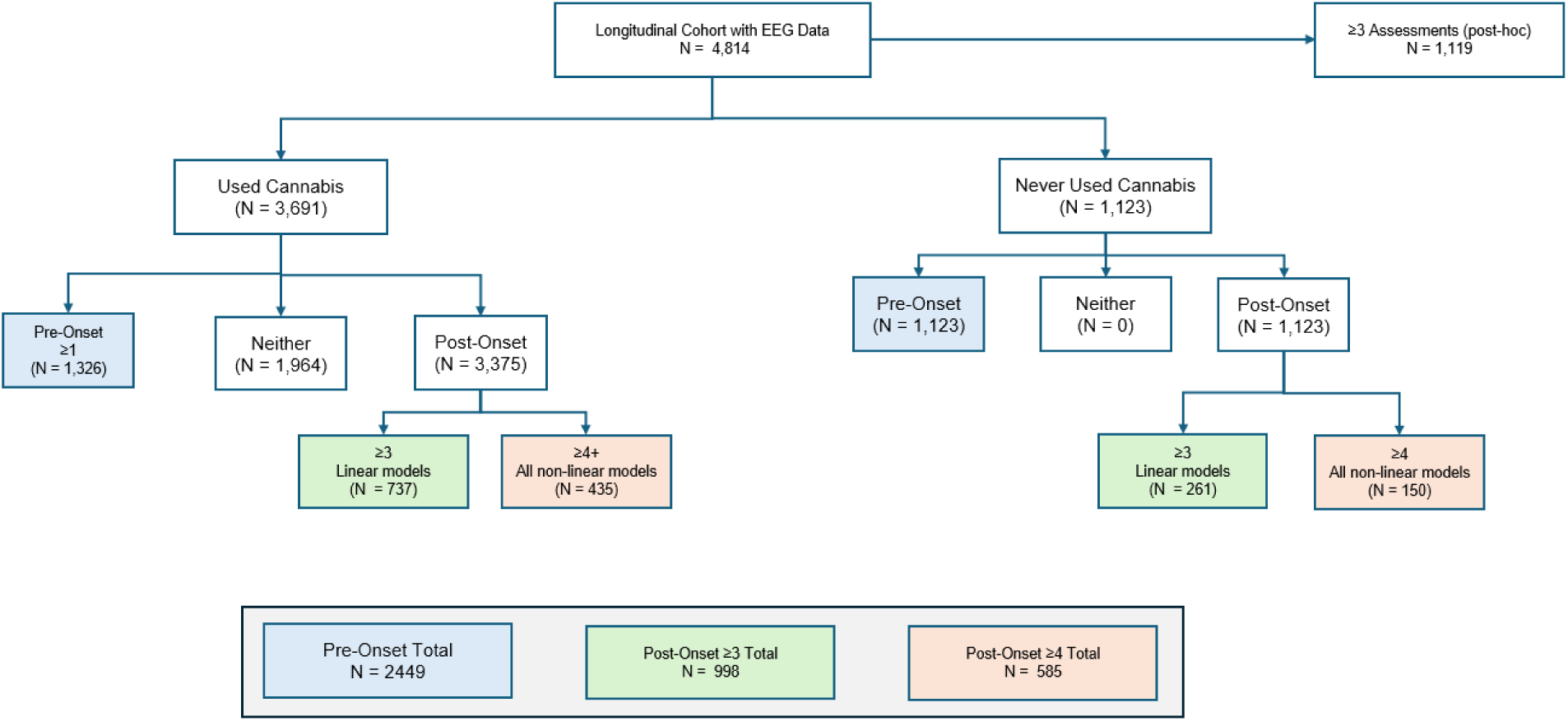
Flow chart of analytic subsamples of COGA participants Flow chart of analytic subsamples of COGA participants split by pre-onset cannabis use. Shaded boxes indicate participants included in the analytic sample totals. Blue denotes the pre-onset group, green denotes the ≥3 post-onset group (linear models), and orange denotes the ≥4 post-onset group (all non-linear models).

## Measures

### Visual Oddball Paradigm (VOP)

Briefly, COGA participants were presented with three sets of visual stimuli, including target (letter X; displayed 12.5% of the time), nontarget (squares; 75%), and novel (colored geometric figures and polygons; 12.5%). Participants were instructed to click a button in response to the target stimuli. Full details regarding the task, preprocessing, and extraction of data are in related publications.^34,57^ **Performance Data.** Target trial (X) accuracy (%) and reaction time (ms; correct trials) were acquired. **P3 Data.** P3 peak amplitude (μV; relative to 0-187 pre-stimulus baseline) and latencies (ms; time to maximal positive deflection in 300-700 ms post-stimulus window) in response to target stimuli were measured from the Pz (mid-parietal) electrode lead.

### Cannabis Use

Lifetime cannabis use was derived from the SSAGA question “Have you ever used marijuana or hashish?”, along with the age at which the individual recalled first using cannabis as their age of onset. Age of onset was derived from the first interview at which cannabis use was reported. Follow-up analyses included self-report of cannabis progression, measured by cannabis use (i.e., ≥10 times during the lifetime, frequency during the heaviest use period), lifetime DSM-5 cannabis use disorder (CanUD) diagnosis)^58^ and severity (i.e., count of the 11 DSM-5 criteria) at last available assessment.

### Covariates

Covariates included sex,^10,59,60^ regular alcohol drinking^61^ (i.e., ever drinking once per month for ≥6 months), cigarette smoking^62^ (i.e., ever smoking a full cigarette), and case status (i.e., relative of original cohort member who was in treatment for AUD during initial ascertainment). Age at P3 assessment was included in survival models, whereas baseline age was included in trajectory models. Regular alcohol use and cigarette smoking were modeled as time-varying covariates in survival analyses and stable covariates in trajectory models. Additional psychiatric and family history covariates (e.g., MDD, ASPD, EXT, INT, parental CanUD) did not meaningfully affect results and were excluded.

### Statistical Methods

#### Pre-Onset analysis

Due to the limited number of individuals reporting lifetime cannabis use with ≥3 assessments before initiation, latent growth curve modeling was not feasible. Instead, Cox proportional hazard models (stcox, STATA SE 18.5) estimated the influence of prior performance and P3 activity on cannabis initiation. Predictors were standardized (mean=0, SD=1) and false discovery rate (FDR) corrections applied. Proportional hazard violations were tested with the Grambsch-Therneau test and corrected for using age interactions (e.g., age ≥18). The Huber-White robust variance estimator accounted for family clustering. Sex interactions were tested but were not significant and are not reported.

#### Post-Onset analysis

Latent growth curve models were fitted in MPlus version 9 to estimate intercepts and slopes for performance and P3 measures. Models were estimated using a robust estimator to account for family clustering. Growth was defined using age in 2-year intervals, with the intercept set at 12-13 years. Raw data were plotted to determine the optimal growth form (linear, quadratic, or piecewise slope). After optimizing model fit (comparative fit index (CFI)>0.95 and the root mean square error of approximation (RMSEA)<0.06), intercepts and slopes were regressed on cannabis initiation and relevant covariates. Although sex interactions were not significant, sex stratified analyses were conducted to account for known sex differences in P3 neurodevelopmental.^10,59,60^ Resulting slopes were extracted and used to estimate odds ratios (O.R.) for cannabis progression outcomes (i.e., ≥10 lifetime uses, frequency of use during heaviest period, CanUD diagnosis and severity) in Stata, applying FDR corrections. In complementary analyses, cannabis use was modeled as a time-varying predictor to capture within person cognitive changes associated with onset.

## Results

### Sample Characteristics

Participants (N=4814) included 52% females, 68% individuals who self-identified as White, and 79% from proband families (**Table 1**). Most participants reported lifetime cannabis use (77%, mean age of onset=17.57, SD=6.09). Regular alcohol use (83%), and cigarette smoking (64%) were common. As the Pre-onset subsample required at ≥1 P3 assessment prior to cannabis onset, participants were younger at first and last assessments but had a later mean age of cannabis initiation than the Post-onset subsample. As expected, lifetime prevalence of cannabis use, cigarette smoking, and regular alcohol use was higher in the Post-onset than Pre-onset subsample. Cannabis use at first assessment was unrelated to participation in ≥3 subsequent follow-up visits (O.R.=1.14, p=0.16).

**Table 1.**
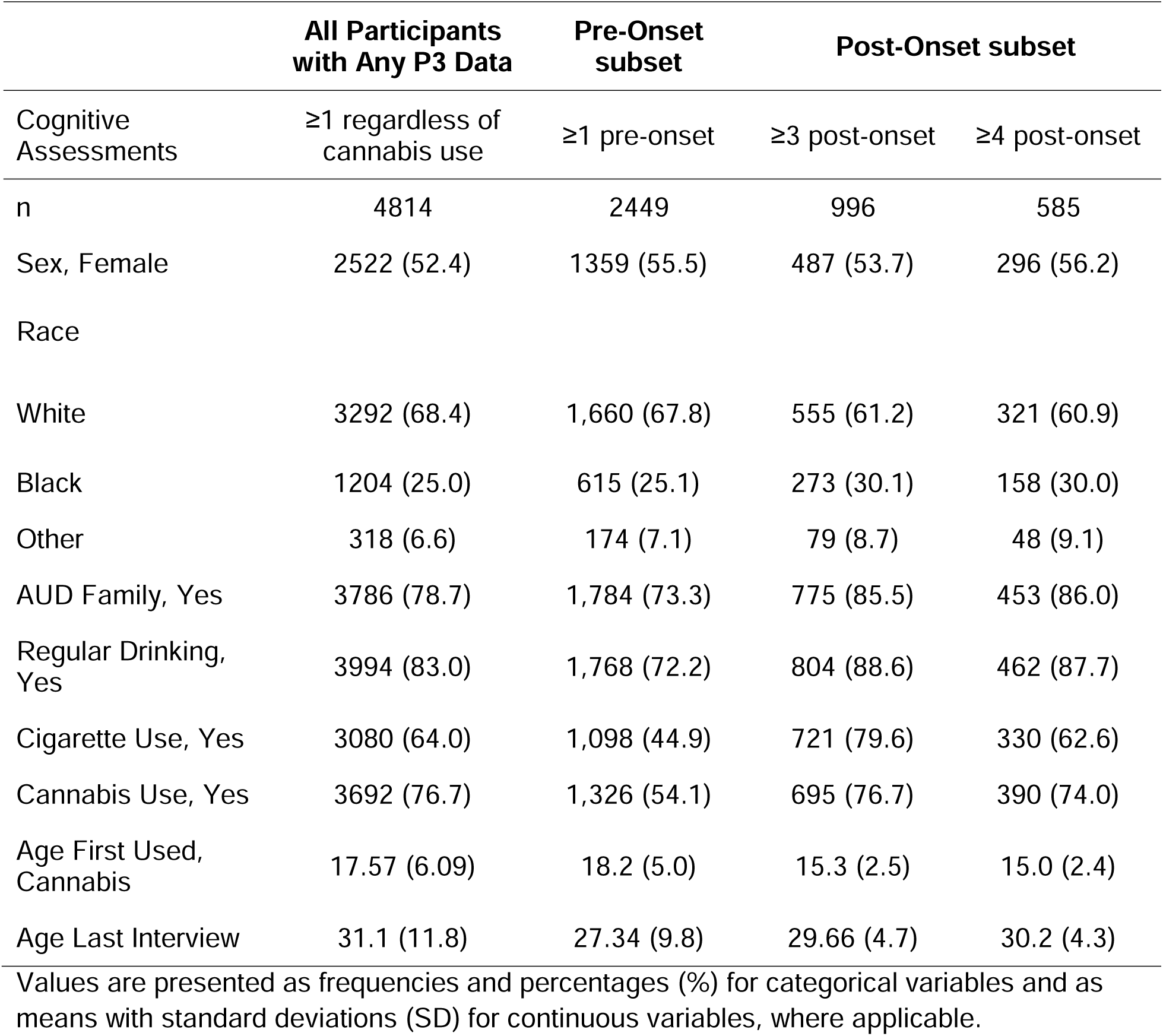
Characteristics of the COGA Prospective-Lifespan participants who have P3 Data.

### Pre-onset subset

#### Does baseline cognition correlate with cannabis initiation

High P3 amplitude was associated with a reduced hazard of cannabis initiation (**Figure 2**), H.R.=.91, 95%CI[.85,.97], p=.006). Prolonged P3 latency and reaction time were associated with a reduced hazard of cannabis initiation. However, proportional hazards testing indicated these effects were only evident when initiation occurred at age ≥18 (H.R._Lat_=.72, 95%CI[.65,.81], p<.001; H.R._RT_=.84, 95%CI[.75,.95],p=.007). Accuracy was not associated with initiation (p=.893).

**Figure 2.**
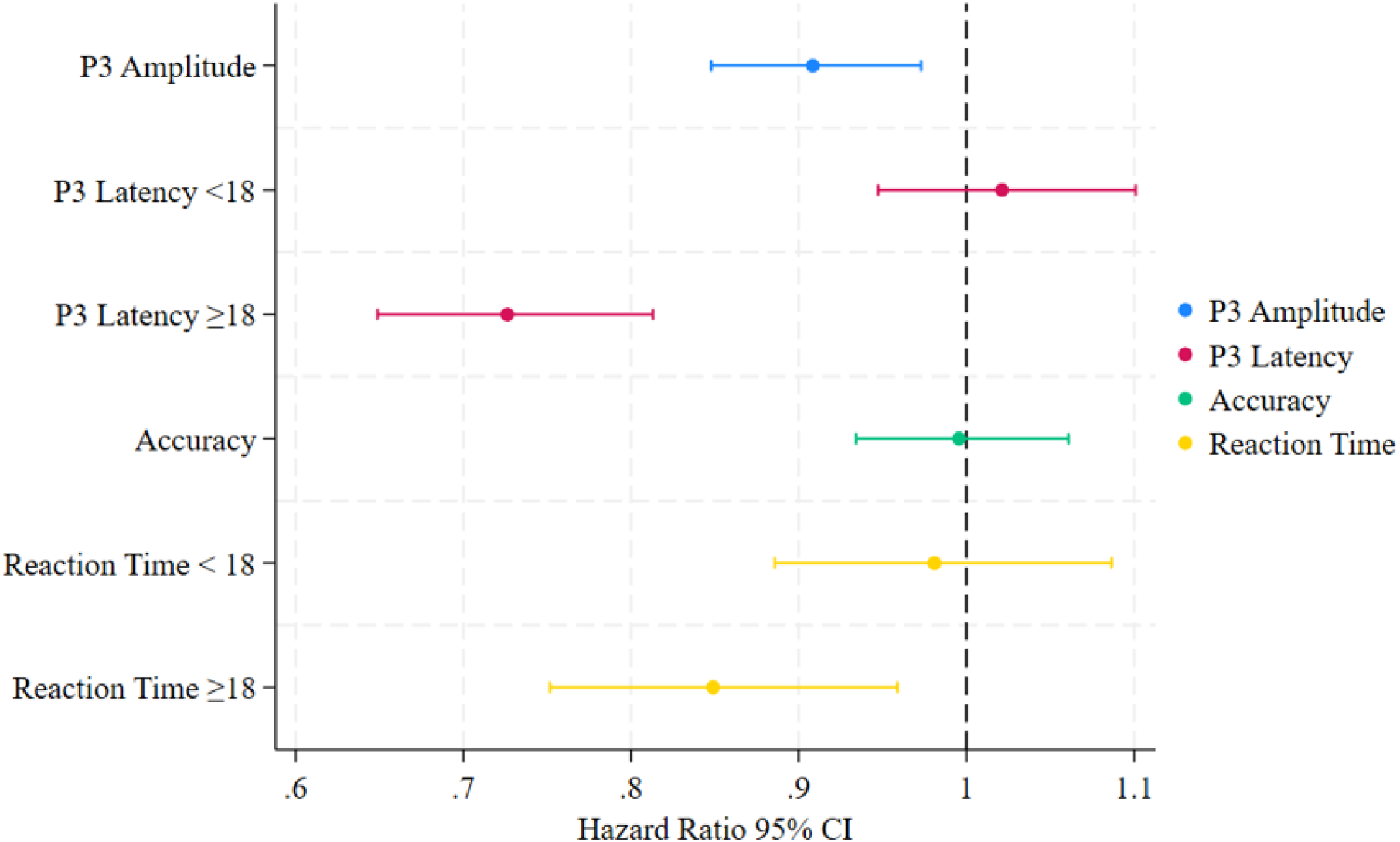
Pre-Onset Hazard Ratios of Cannabis Initiation Onset Hazard Ratios [HR] with 95% confidence intervals (CIs) for cannabis initiation in the pre-onset subsample. Latency and reaction time models are presented separately for onset before age 18 (< 18) and age 18 and older (≥18) to address violations of the proportional hazard’s assumption.

### Post-onset subset

#### Does cannabis initiation relate to subsequent cognitive trajectories

##### P3 amplitude

P3 amplitudes decreased across age intervals, supporting a significant linear slope (**Figure 3**; mean slope: b=-1.54, 95%C.I.[-2.02,-1.06]; p<.001; CFI=.98; RMSEA=0.026]. Cannabis initiation was not associated with P3 amplitude at age 12-13 (i.e., intercept: b=.03,95%C.I.[-0.08,0.15], p=0.64). However, initiation was associated with a significantly steeper decline in P3 amplitude in the combined (mean slope: b=-0.29, 95%C.I.[-0.50,-0.09]; p=0.002), and female stratified (b=-0.58, 95%C.I.[-0.91,-0.25]; p=0.004; **eFigure 1**) samples, but not in males (b=0.004,95%C.I.[-0.28,0.29]; p=0.98). Intercepts remained uncorrelated in both sexes. In complementary time-varying analyses, initiation was not associated with P3 amplitude at ages ≤20 (all p’s>.05) but was associated with lower P3 amplitude at ages ≥20 (range of b’s=-0.04 to −0.12, range of p’s=.025 to <.001; **eTable 1**).

**Figure 3.**
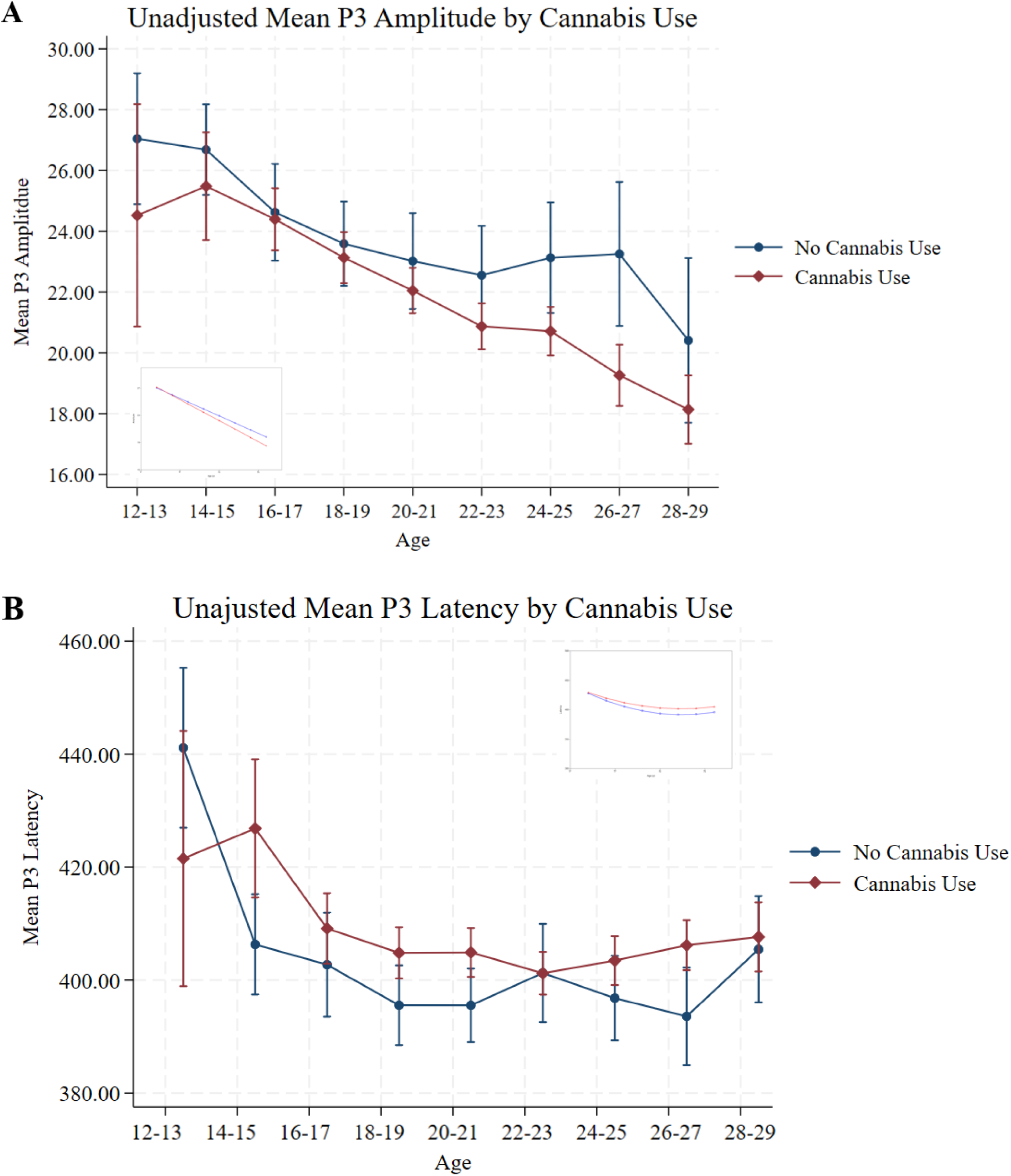
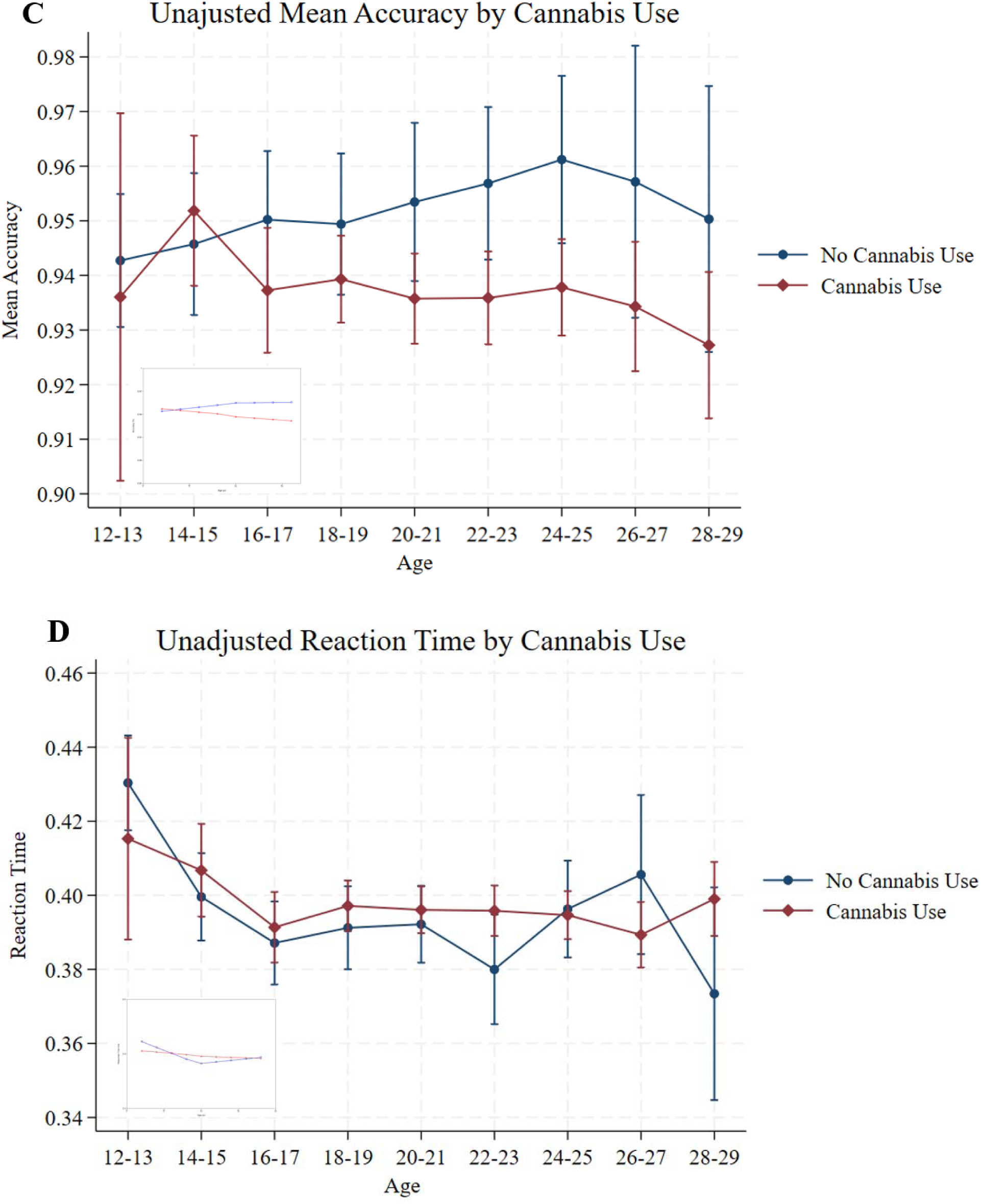
Post-Onset P3 and Performance Trajectories by Cannabis Use Unadjusted mean changes in P3 amplitude (A) and latency (B), and Visual Oddball (VO) task accuracy (C) and response time (D) from ages 12 to 29 by cannabis use. Error bars represent 95% confidence intervals (CI). The inset panels display the selected latent growth curve adjusted for covariates.

##### P3 latency

P3 latency suggests a U-shaped distribution, decreasing at ages <18 and increasing at ages ≥18. Consistent with this, both linear (mean slope: b=-10.45, 95%C.I.[-12.97,-7.92];p<.001) and quadratic [b=1.02, 95% C.I.[.75,1.29], p<.001) growth parameters were significant. Model fit improved with the inclusion of the quadratic term (CFI = .98; RMSEA=0.017) compared to a linear only model (CFI=.87; RMSEA =0.037). Cannabis initiation was associated with the intercept or growth parameter regardless of sex (all p’s>.05).

##### Accuracy

Accuracy remained high across age intervals (range: 93-95%) and the linear growth estimate was not significant (mean slope: b=-0.020, 95%C.I.[-0.32,0.28], p=0.912; CFI=.93; RMSEA=0.030).

##### Reaction time

Reaction time slightly decreased linearly until age 18, then stabilized (range: .42-.39 ms). Linear (mean slope: b=-0.26, 95%C.I.[-0.36,-0.16]; p<.001) and piecewise (b_<18_=0.44, 95%C.I.[-0.61,-0.27], p<.001; b_≥18_=-0.06, 95%C.I.[-0.21,0.09], p=.50) growth parameters were significant. Model fit was improved using the piecewise model (CFI=.94; RMSEA=0.037) compared to the linear (CFI=.92; RMSEA=0.035) model. Cannabis initiation was associated with faster reaction time at age 12-13 (i.e., intercept: b=-0.27, 95%CI[-0.40,-0.14],p<.001). Initiation was also associated with stabilized reaction time, relative to the steeper declines observed in the those who did not initiate cannabis use, in the combined (mean slope: b=0.35, 95%C.I.[0.15,0.55]; p=0.005), and female stratified (b=0.48, 95%C.I.[0.20,0.77];p=0.006) samples, but not in males (b= 0.14, 95%C.I.[-0.18,0.46];p=0.46) until age 18, after which trajectories did not vary (all p’s>.05). In complementary time-varying analyses, cannabis initiation was consistently associated with slower reaction time at ages ≥22 (range of b’s=0.04 to 0.10, range of p’s=.002 to .048).

#### Does cognitive change correlate with odds of cannabis progression

Post-hoc associations between cannabis progression measures and growth model slope estimates for reaction time and P3 amplitude (i.e., cognition changes associated with initiation) documented that steeper decline in P3 amplitude was associated with greater odds of self-report of cannabis use ≥10 times (O.R.=2.33,p<.001), frequency of use during heaviest period (β=0.16,p=.001), CanUD diagnosis (O.R.=2.34,p<.001) and severity, as indexed by a criterion count (β=.31,p<.001), while steeper decrease in reaction time was associated with reduced odds of cannabis use ≥10 times (O.R.=.60,p<.001), CanUD diagnosis (O.R.=.79,p=.002), and severity (β=-.08, p=.018; **Figure 4**)

**Figure 4.**
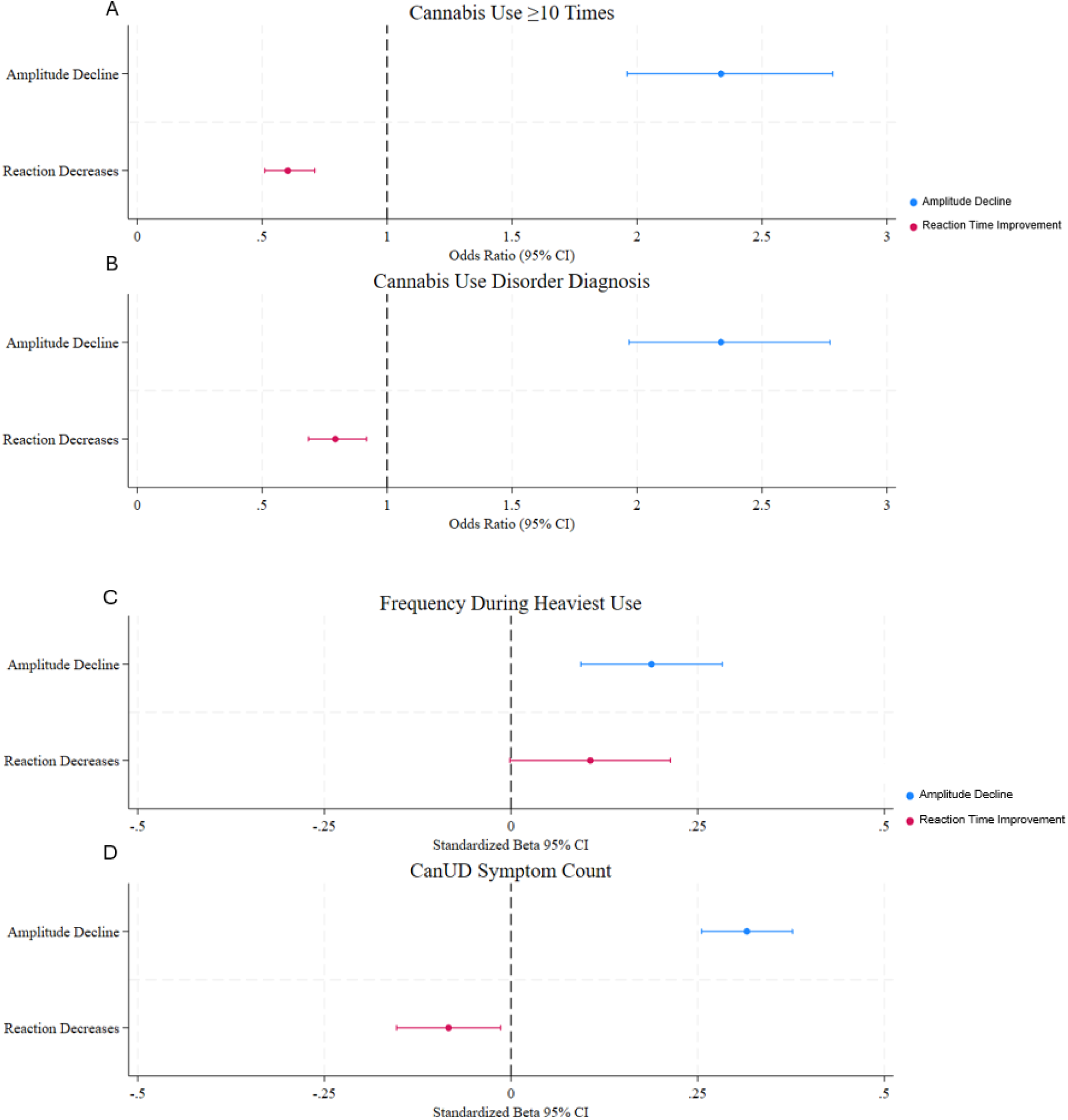
Associations Between Cannabis Progression Measures and Growth Model Slope Estimates Odds Ratios (OR) and β with 95% confidence intervals (CIs) for self-reported cannabis is use (≥10 times during the lifetime, frequency of use during heaviest period), DSM-5 cannabis use disorder, and severity (i.e., symptom count) in the post-onset subsample. Amplitude and Response Time have been reverse-coded for to represent steeper declines (i.e. reaction decreases).

## Discussion

Cannabis use is frequently linked to changes in cognition,^10–15^ but few studies have collected longitudinal neurocognitive data before and after initiation to distinguish preexisting from cannabis-related differences. Such distinctions are important given cannabis initiation often coincides with neurodevelopment.^26,27^ Clarifying these temporal relationships may help distinguish putatively “causal” effects of cannabis use,^29^ the neurological mechanisms involved, and ultimately their associations with broader psychosocial outcomes. The present study differentiated preexisting and onset-subsequent cognitive differences associated with cannabis use and examined whether cannabis-related cognitive changes were more likely to correlate with cannabis use progression.

Several notable findings emerged. First, high P3 amplitude and prolonged P3 latency and reaction time were associated with a decreased hazard of cannabis initiation. Second, individuals who initiated cannabis use showed steeper subsequent declines in P3 amplitude, occurring alongside faster but stabilized response times. In contrast, those without a history of cannabis use evidenced age-related decreases in amplitude and maintained accuracy while demonstrating reductions in response times, consistent with improved efficiency with practice. Together, this findings suggest differences in brain maturation may both increase risk for cannabis initiation and contribute to cognitive differences that may otherwise be misinterpreted solely as causal effects of cannabis exposure.^63^

P3 amplitude typically peaks during adolescence (∼16 years) and subsequently decreases throughout young adulthood, while P3 latency generally decreases until early adulthood (∼22 years), reflecting increasing neural efficiency and faster cognitive processing.^64–66^ Therefore, the steeper P3 amplitude declines and relatively earlier optimized reaction times observed among those who initiated cannabis use may indicate accelerated neurodevelopmental trajectories, particularly given similarly high accuracy rates across groups. Furthermore, the high baseline P3 amplitude and prolonged P3 latency and reaction time associated with cannabis initiation may suggest that individuals who do not initiate cannabis use are “earlier”, or less mature in their neurodevelopmental trajectories before exposure. These findings align with early work that suggesting substance use onset may be associated with accelerated developmental trajectories.^67,68^ For instance, one longitudinal study found that youth who used cannabis initially demonstrated relatively better cognitive performance (e.g., working memory, inhibitory control, visuospatial processing) during late childhood, but later displayed flattened trajectories over time.^63^ Alcohol research has similarly demonstrated accelerated neurodevelopmental trajectories among heavy drinking youth, including decreased cortical thickness and accelerated gray matter volume decline.^69–72^

Baseline P3 latency and reaction time were also only statistically significant factors for adult-onset cannabis use (≥18 years), with no influence of prior cognition on earlier onset. Cross-sectional analyses of the time-varying influence of cannabis use on P3 amplitude and reaction time similarly identified significant associations after adulthood (≥20 and ≥22 years, respectively). Given the temporality of normal P3 amplitude and latency development,^64–66^ these findings may suggest that accelerated neurodevelopment associated with cannabis initiation require time to emerge behaviorally and neurophysiologically, becoming more salient over time. Additionally, although cognitive changes following cannabis initiation were observed in the combined sample, stratified models suggest that effects were largely driven by females, consistent with prior work. Estrogen is a well-documented modulator of the endocannabinoid system^73,74^ potentially amplifying cannabis’ effects. Sex-dependent differences in THC metabolism have also been observed in preclinical studies^75^. Additionally, sex-specific differences in developmental trajectories of P3 have also been observed^76–78^, and females generally exhibit earlier cortical neural maturation^79,80^ and greater declines in P3 trajectories.^10,59,60^

Lastly, steeper P3 amplitude decreases, and stabilized reaction times reflected a deepening and problematic engagement with cannabis, including heavier, more frequent use and CanUD, assessed towards the end of the “growth” period. These findings are consistent with evidence that suggests cannabis use may exert a potentially neurotoxic and dose-responsive influence on measures of cognition,^11,13,16–18^ including P3. Prior studies have found lagged within-subject effects of increasing cannabis involvement on cognitive performance tasks^16^ and reduced activation during a working memory task in addiction-relevant brain regions (e.g., insula, medial and dorsolateral prefrontal cortex) in individuals using cannabis heavily.^14^ Reduced P3 amplitude has also been observed among individuals using cannabis and following THC administration,^10,81–86^ with one study reporting dose related reductions in P3 amplitude post-THC infusion changes in P3.^82^ However, it is not possible given the current study design to make definitive causal inferences, nor confirm or refute a neurotoxic effect of cannabis use on cognition. Indeed, pleiotropic effects of genes or “third” variables, such as early life environments may jointly contribute to cognitive variability and to likelihood of problematic cannabis use. However, taken together, the current study findings suggest that cognitive differences associated with use reflect both preexisting developmental variation and are related to cannabis exposure.

Some limitations warrant consideration. First, despite the longitudinal design, causal conclusions cannot be made, as additional factors may contribute to both cognitive functioning and cannabis use. However, findings remained consistent in sensitivity analyses accounting for family history of cannabis use and other psychiatric disorders. Second, as COGA is enriched for alcohol use disorder, results may not generalize to other less selected cohorts. However, our results broadly align with those from other cohort and experimental studies.^67,68^ Third, attrition may have introduced bias, with individuals with greater cannabis use or poorer neurocognition less likely to return for multiple assessments. Some reassurance is gained from noting that cross-sectional analyses that did not rely on longitudinal retention recapitulated the main findings (**eTable 1**) and cannabis use at first assessment was unrelated to participation at ≥3 subsequent follow-up visits.

## Conclusion

These findings indicate that accelerated neurodevelopment may modify the likelihood of cannabis initiation which, in turn, may further contribute to neurocognitive changes that deepen cannabis involvement. This work highlights the importance of longitudinal approaches capable of disentangling predispositional from cannabis-related cognitive differences and suggests that accelerated neurodevelopmental trajectories may represent one pathway linking cognition and cannabis use across adolescence and young adulthood.

## Data Availability

COGA data access is restricted but can be accessed via an application to the National Institute on Alcohol Abuse and Alcoholism or in collaboration with a COGA investigator as a sponsor for a secondary analysis proposal. Details regarding access at https://cogastudy.org/resources-for-researchers/#accessing-coga-data.

## Acknowledgements

The Collaborative Study on the Genetics of Alcoholism (COGA), Principal Investigators B. Porjesz, V. Hesselbrock, A. Agrawal; Scientific Director, A. Agrawal; Translational Director, D. Dick, includes ten different centers: University of Connecticut (V. Hesselbrock); Indiana University (H.J. Edenberg, T. Foroud, Y. Liu, M.H. Plawecki); University of Iowa Carver College of Medicine (S. Kuperman, A. Anderson); SUNY Downstate Health Sciences University (B. Porjesz, J. Meyers); Washington University in St. Louis (L. Bierut, A. Agrawal, S. Hartz); University of California at San Diego (M. Schuckit); Rutgers University (D. Dick, R. Hart, J. Salvatore, J. Tischfield); The Children’s Hospital of Philadelphia, University of Pennsylvania (L. Almasy); Icahn School of Medicine at Mount Sinai (A. Goate, P. Slesinger); and Howard University (D. Scott). Other COGA collaborators include: C. Holzhauer, M. Hesselbrock (University of Connecticut); D. Lai, J. Nurnberger Jr., L. Wetherill, X., Xuei, S. O’Connor, (Indiana University); J. Kramer (University of Iowa), G. Chan (University of Iowa; University of Connecticut); C. Kamarajan, A. Pandey, D.B. Chorlian, P. Barr, S. Kinreich, G. Pandey, Z. Neale, C. Chatzinakos, J. Zhang, S. Saenz deViteri, R. Christian, A. Bingly (SUNY Downstate); G. Pathak (Icahn School of Medicine at Mount Sinai); A. Anokhin, K. Bucholz, F. Dong, A. Hatoum, E. Johnson, V. McCutcheon, J. Rice, S. Saccone (Washington University); F. Aliev, Z. Pang, S. Kuo, S. Brislin, J. Moore (Rutgers University); A. Merikangas (The Children’s Hospital of Philadelphia and University of Pennsylvania); M. Gitik, NIAAA Staff Collaborator. We continue to be inspired by our memories of Henri Begleiter and Theodore Reich, founding PI and Co-PI of COGA, and also owe a debt of gratitude to other past organizers of COGA, including Ting-Kai Li, P. Michael Conneally, Raymond Crowe, and Wendy Reich, for their critical contributions. This national collaborative study is supported by NIH Grant U10AA008401 from the National Institute on Alcohol Abuse and Alcoholism (NIAAA) and the National Institute on Drug Abuse (NIDA).

## References

1. Miech RA, Johnston LD, Patrick ME, O’Malley PM. Monitoring the Future National Survey Results on Drug Use, 1975-2024: Overview and Detailed Results for Secondary School Students. Monitoring the Future Monograph Series. Institute for Social Research, University of Michigan; 2025.

2. Miech RA, Johnston LD, Patrick ME, O’Malley PM, Bachman JG. Monitoring the Future National Survey Results on Drug Use, 1975-2023: Overview and Detailed Results for Secondary School Students. Institute for Social Research; 2024.

3. Volkow ND, Baler RD, Compton WM, Weiss SRB. Adverse Health Effects of Marijuana Use. N Engl J Med. 2014;370(23):2219–2227. doi:10.1056/NEJMra1402309

4. Silins E, Horwood LJ, Patton GC, et al. Young adult sequelae of adolescent cannabis use: an integrative analysis. Lancet Psychiatry. 2014;1(4):286–293. doi:10.1016/S2215-0366(14)70307-4

5. Marie O, Zölitz U. ‘High’ Achievers? Cannabis Access and Academic Performance. Rev Econ Stud. Published online March 27, 2017. doi:10.1093/restud/rdx020

6. Zalesky A, Solowij N, Yucel M, et al. Effect of long-term cannabis use on axonal fibre connectivity. Brain. 2012;135(7):2245–2255. doi:10.1093/brain/aws136

7. Dahlgren MK, Sagar KA, Smith RT, Lambros AM, Kuppe MK, Gruber SA. Recreational cannabis use impairs driving performance in the absence of acute intoxication. Drug Alcohol Depend. 2020;208:107771. doi:10.1016/j.drugalcdep.2019.107771

8. Chadwick B, Miller ML, Hurd YL. Cannabis Use during Adolescent Development: Susceptibility to Psychiatric Illness. Front Psychiatry. 2013;4. doi:10.3389/fpsyt.2013.00129

9. Urits I, Gress K, Charipova K, et al. Cannabis Use and its Association with Psychological Disorders. Psychopharmacol Bull. 2025;50(2):56–67. doi:10.64719/pb.4606

10. Schiemer C, Summers MJ, Stefanidis KB. Identifying EEG markers related to acute cannabis consumption: A systematic review. Neurosci Biobehav Rev. 2025;172:106092. doi:10.1016/j.neubiorev.2025.106092

11. Kroon E, Kuhns L, Cousijn J. The short-term and long-term effects of cannabis on cognition: recent advances in the field. Curr Opin Psychol. 2021;38:49–55. doi:10.1016/j.copsyc.2020.07.005

12. Dellazizzo L, Potvin S, Giguère S, Dumais A. Evidence on the acute and residual neurocognitive effects of cannabis use in adolescents and adults: a systematic meta-review of meta-analyses. Addiction. 2022;117(7):1857–1870. doi:10.1111/add.15764

13. Lorenzetti V, Hoch E, Hall W. Adolescent cannabis use, cognition, brain health and educational outcomes: A review of the evidence. Eur Neuropsychopharmacol. 2020;36:169–180. doi:10.1016/j.euroneuro.2020.03.012

14. Gowin JL, Ellingson JM, Karoly HC, et al. Brain Function Outcomes of Recent and Lifetime Cannabis Use. JAMA Netw Open. 2025;8(1):e2457069. doi:10.1001/jamanetworkopen.2024.57069

15. Debenham J, Birrell L, Champion K, Lees B, Yücel M, Newton N. Neuropsychological and neurophysiological predictors and consequences of cannabis and illicit substance use during neurodevelopment: a systematic review of longitudinal studies. Lancet Child Adolesc Health. 2021;5(8):589–604. doi:10.1016/S2352-4642(21)00051-1

16. Morin JFG, Afzali MH, Bourque J, et al. A Population-Based Analysis of the Relationship Between Substance Use and Adolescent Cognitive Development. Am J Psychiatry. 2019;176(2):98–106. doi:10.1176/appi.ajp.2018.18020202

17. Morie KP, Potenza MN. A Mini-Review of Relationships Between Cannabis Use and Neural Foundations of Reward Processing, Inhibitory Control and Working Memory. Front Psychiatry. 2021;12:657371. doi:10.3389/fpsyt.2021.657371

18. Tervo-Clemmens B, Simmonds D, Calabro FJ, Day NL, Richardson GA, Luna B. Adolescent cannabis use and brain systems supporting adult working memory encoding, maintenance, and retrieval. NeuroImage. 2018;169:496–509. doi:10.1016/j.neuroimage.2017.12.041

19. Fergusson DM, Boden JM. Cannabis use and later life outcomes. Addiction. 2008;103(6):969–976. doi:10.1111/j.1360-0443.2008.02221.x

20. Patrick ME, Pang YC, Terry-McElrath YM, Jang JB. Young Adult Substance Use as a Predictor of Poor Self-Rated Memory Decades Later in Midlife. J Aging Health. Published online March 4, 2026:08982643261431007. doi:10.1177/08982643261431007

21. Meyer HC, Lee FS, Gee DG. The Role of the Endocannabinoid System and Genetic Variation in Adolescent Brain Development. Neuropsychopharmacology. 2018;43(1):21–33. doi:10.1038/npp.2017.143

22. Harkany T, Guzmán M, Galve-Roperh I, Berghuis P, Devi LA, Mackie K. The emerging functions of endocannabinoid signaling during CNS development. Trends Pharmacol Sci. 2007;28(2):83–92. doi:10.1016/j.tips.2006.12.004

23. Durieux LJA, Gilissen SRJ, Arckens L. Endocannabinoids and cortical plasticity: CB1R as a possible regulator of the excitation/inhibition balance in health and disease. Eur J Neurosci. 2022;55(4):971–988. doi:10.1111/ejn.15110

24. Cannabis exposure during adolescence: A uniquely sensitive period for neurobiological effects. In: International Review of Neurobiology. Vol 161. Elsevier; 2021:95–120. doi:10.1016/bs.irn.2021.07.002

25. Fischer AS, Tapert SF, Louie DL, Schatzberg AF, Singh MK. Cannabis and the Developing Adolescent Brain. Curr Treat Options Psychiatry. 2020;7(2):144–161. doi:10.1007/s40501-020-00202-2

26. Schneider M. Puberty as a highly vulnerable developmental period for the consequences of cannabis exposure. Addict Biol. 2008;13(2):253–263. doi:10.1111/j.1369-1600.2008.00110.x

27. Hall WD, Patton G, Stockings E, et al. Why young people’s substance use matters for global health. Lancet Psychiatry. 2016;3(3):265–279. doi:10.1016/S2215-0366(16)00013-4

28. Uhl GR, Koob GF, Cable J. The neurobiology of addiction. Ann N Y Acad Sci. 2019;1451(1):5–28. doi:10.1111/nyas.13989

29. Jacobus J, Tapert S. Effects of Cannabis on the Adolescent Brain. Curr Pharm Des. 2014;20(13):2186–2193. doi:10.2174/13816128113199990426

30. Nguyen-Louie TT, Castro N, Matt GE, Squeglia LM, Brumback T, Tapert SF. Effects of Emerging Alcohol and Marijuana Use Behaviors on Adolescents’ Neuropsychological Functioning Over Four Years. J Stud Alcohol Drugs. 2015;76(5):738–748. doi:10.15288/jsad.2015.76.738

31. Jacobus J, Squeglia LM, Infante MA, et al. Neuropsychological performance in adolescent marijuana users with co-occurring alcohol use: A three-year longitudinal study. Neuropsychology. 2015;29(6):829–843. doi:10.1037/neu0000203

32. Becker MP, Collins PF, Schultz A, Urošević S, Schmaling B, Luciana M. Longitudinal changes in cognition in young adult cannabis users. J Clin Exp Neuropsychol. 2018;40(6):529–543. doi:10.1080/13803395.2017.1385729

33. Meier MH, Caspi A, Ambler A, et al. Persistent cannabis users show neuropsychological decline from childhood to midlife. Proc Natl Acad Sci. 2012;109(40). doi:10.1073/pnas.1206820109

34. Cohen HL, Wang W, Porjesz B, et al. Visual P300: An interlaboratory consistency study. Alcohol. 1994;11(6):583–587. doi:10.1016/0741-8329(94)90087-6

35. Polich J. P300 as a clinical assay: rationale, evaluation, and findings. Int J Psychophysiol. 2000;38(1):3–19. doi:10.1016/S0167-8760(00)00127-6

36. Luck, S.J. An Introduction to the Event-Related Potential Technique; MIT Press: Cambridge, MA USA, 2014.

37. Bewernitz M, Derendorf H. Electroencephalogram-based pharmacodynamic measures: a review. Int J Clin Pharmacol Ther. 2012;50(03):162–184. doi:10.5414/CP201484

38. Volpe U, Mucci A, Bucci P, Merlotti E, Galderisi S, Maj M. The cortical generators of P3a and P3b: A LORETA study. Brain Res Bull. 2007;73(4-6):220–230. doi:10.1016/j.brainresbull.2007.03.003

39. Goldstein A, Spencer KM, Donchin E. The influence of stimulus deviance and novelty on the P300 and Novelty P3. Psychophysiology. 2002;39(6):781–790. doi:10.1111/1469-8986.3960781

40. Katayama J, Polich J. Stimulus context determines P3a and P3b. Psychophysiology. 1998;35(1):23–33. doi:10.1111/1469-8986.3510023

41. Falkenstein M, Hoormann J, Hohnsbein J. ERP components in Go/Nogo tasks and their relation to inhibition. Acta Psychol (Amst). 1999;101(2-3):267–291. doi:10.1016/S0001-6918(99)00008-6

42. Polich J. Updating P300: An integrative theory of P3a and P3b. Clin Neurophysiol. 2007;118(10):2128–2148. doi:10.1016/j.clinph.2007.04.019

43. Portin R, Kovala T, Polo-Kantola P, Revonsuo A, Müller K, Matikainen E. Does P3 Reflect Attentional or Memory Performances, or Cognition more Generally? Scand J Psychol. 2000;41(1):31–40. doi:10.1111/1467-9450.00168

44. Reed CL, Siqi-Liu A, Lydic K, et al. Selective contributions of executive function ability to the P3. Int J Psychophysiol. 2022;176:54–61. doi:10.1016/j.ijpsycho.2022.03.004

45. Polich J. Neuropsychology of P300. Oxford University Press; 2011. doi:10.1093/oxfordhb/9780195374148.013.0089

46. Spronk DB, De Bruijn ERA, Van Wel JHP, Ramaekers JG, Verkes RJ. Acute effects of cocaine and cannabis on response inhibition in humans: an ERP investigation: Drugs on response inhibition. Addict Biol. 2016;21(6):1186–1198. doi:10.1111/adb.12274

47. Böcker KBE, Gerritsen J, Hunault CC, Kruidenier M, Mensinga TjT, Kenemans JL. Cannabis with high Δ9-THC contents affects perception and visual selective attention acutely: An event-related potential study. Pharmacol Biochem Behav. 2010;96(1):67–74. doi:10.1016/j.pbb.2010.04.008

48. Hart CL, Ilan AB, Gevins A, et al. Neurophysiological and cognitive effects of smoked marijuana in frequent users. Pharmacol Biochem Behav. 2010;96(3):333–341. doi:10.1016/j.pbb.2010.06.003

49. Richard CD, Poole JR, McConnell M, et al. Alterations in Electroencephalography Theta as Candidate Biomarkers of Acute Cannabis Intoxication. Front Neurosci. 2021;15:744762. doi:10.3389/fnins.2021.744762

50. Iacono WG, Carlson SR, Malone SM, McGue M. P3 Event-Related Potential Amplitude and the Risk for Disinhibitory Disorders in Adolescent Boys. Arch Gen Psychiatry. 2002;59(8):750. doi:10.1001/archpsyc.59.8.750

51. Harper J, Malone SM, Iacono WG. Parietal P3 and midfrontal theta prospectively predict the development of adolescent alcohol use. Psychol Med. 2021;51(3):416–425. doi:10.1017/S0033291719003258

52. Agrawal A, Brislin SJ, Bucholz KK, et al. The Collaborative Study on the Genetics of Alcoholism: Overview. Genes Brain Behav. 2023;22(5):e12864. doi:10.1111/gbb.12864

53. The Collaborative Study on the Genetics of Alcoholism. Alcohol Health Res World. 1995;19(3):228–236.

54. Bucholz KK, McCutcheon VV, Agrawal A, et al. Comparison of Parent, Peer, Psychiatric, and Cannabis Use Influences Across Stages of Offspring Alcohol Involvement: Evidence from the COGA Prospective Study. Alcohol Clin Exp Res. 2017;41(2):359–368. doi:10.1111/acer.13293

55. Bucholz KK, Cadoret R, Cloninger CR, et al. A new, semi-structured psychiatric interview for use in genetic linkage studies: a report on the reliability of the SSAGA. J Stud Alcohol. 1994;55(2):149–158. doi:10.15288/jsa.1994.55.149

56. Hesselbrock M, Easton C, Bucholz KK, Schuckit M, Hesselbrock V. A validity study of the SSAGA-a comparison with the SCAN. Addiction. 1999;94(9):1361–1370. doi:10.1046/j.1360-0443.1999.94913618.x

57. Porjesz B, Begleiter H, Reich T, et al. Amplitude of Visual P3 Event-Related Potential as a Phenotypic Marker for a Predisposition to Alcoholism: Preliminary Results from the COGA Project. Alcohol Clin Exp Res. 1998;22(6):1317–1323. doi:10.1111/j.1530-0277.1998.tb03914.x

58. American Psychiatric Association. Diagnostic and Statistical Manual of Mental Disorders (DSM-5). Fifth Edition ed. Arlington, VA: American Psychiatric Association; 2013.

59. Sumich AL, Sarkar S, Hermens DF, et al. Sex Differences in Brain Maturation as Measured Using Event-Related Potentials. Dev Neuropsychol. 2012;37(5):415–433. doi:10.1080/87565641.2011.653461

60. Brumback TY, Arbel Y, Donchin E, Goldman MS. Efficiency of responding to unexpected information varies with sex, age, and pubertal development in early adolescence. Psychophysiology. 2012;49(10):1330–1339. doi:10.1111/j.1469-8986.2012.01444.x

61. Wickens CM, Wright M, Mann RE, et al. Separate and combined effects of alcohol and cannabis on mood, subjective experience, cognition and psychomotor performance: A randomized trial. Prog Neuropsychopharmacol Biol Psychiatry. 2022;118:110570. doi:10.1016/j.pnpbp.2022.110570

62. Kroon E, De Bode N, Colyer-Patel K, Hsieh JH, Filbey F, Cousijn J. Cross-sectional and longitudinal associations of cannabis use with cognitive functioning in individuals with a cannabis use disorder: The moderating role of nicotine. Addiction. Published online May 6, 2026:add.70457. doi:10.1111/add.70457

63. Wade NE, Sullivan RM, Wallace AL, et al. Longitudinal neurocognitive trajectories in a large cohort of youth who use cannabis: combining self-report and toxicology. Neuropsychopharmacology. Published online April 20, 2026. doi:10.1038/s41386-026-02395-1

64. Mingils SM, Davies PL, Stephens JA, Gavin WJ. Developmental trends of auditory novelty oddball P3 while accounting for N2 in 7- to 25- year-olds. Psychophysiology. 2023;60(4):e14214. doi:10.1111/psyp.14214

65. Martín LJ, Barajas JJ, Fernández R. Auditory P3 development in childhood. Scand Audiol Suppl. 1988;30:105–109.

66. Van Dinteren R, Arns M, Jongsma MLA, Kessels RPC. P300 Development across the Lifespan: A Systematic Review and Meta-Analysis. Di Russo F, ed. PLoS ONE. 2014;9(2):e87347. doi:10.1371/journal.pone.0087347

67. Miller AP, Baranger DAA, Paul SE, et al. Characteristics Associated With Cannabis Use Initiation by Late Childhood and Early Adolescence in the Adolescent Brain Cognitive Development (ABCD) Study. JAMA Pediatr. 2023;177(8):861. doi:10.1001/jamapediatrics.2023.1801

68. Green R, Wolf BJ, Chen A, et al. Predictors of Substance Use Initiation by Early Adolescence. Am J Psychiatry. 2024;181(5):423–433. doi:10.1176/appi.ajp.20230882

69. Pfefferbaum A, Kwon D, Brumback T, et al. Altered Brain Developmental Trajectories in Adolescents After Initiating Drinking. Am J Psychiatry. 2018;175(4):370–380. doi:10.1176/appi.ajp.2017.17040469

70. Luciana M, Collins PF, Muetzel RL, Lim KO. Effects of alcohol use initiation on brain structure in typically developing adolescents. Am J Drug Alcohol Abuse. 2013;39(6):345–355. doi:10.3109/00952990.2013.837057

71. Squeglia LM, Tapert SF, Sullivan EV, et al. Brain Development in Heavy-Drinking Adolescents. Am J Psychiatry. 2015;172(6):531–542. doi:10.1176/appi.ajp.2015.14101249

72. Lees B, Meredith LR, Kirkland AE, Bryant BE, Squeglia LM. Effect of alcohol use on the adolescent brain and behavior. Pharmacol Biochem Behav. 2020;192:172906. doi:10.1016/j.pbb.2020.172906

73. Vallée M, Vitiello S, Bellocchio L, et al. Pregnenolone Can Protect the Brain from Cannabis Intoxication. Science. 2014;343(6166):94–98. doi:10.1126/science.1243985

74. Hill MN, Karacabeyli ES, Gorzalka BB. Estrogen recruits the endocannabinoid system to modulate emotionality. Psychoneuroendocrinology. 2007;32(4):350–357. doi:10.1016/j.psyneuen.2007.02.003

75. Wiley JL, Burston JJ. Sex differences in Δ9-tetrahydrocannabinol metabolism and in vivo pharmacology following acute and repeated dosing in adolescent rats. Neurosci Lett. 2014;576:51–55. doi:10.1016/j.neulet.2014.05.057

76. Hill SY, Jones BL, Steinhauer SR, Zezza N, Stiffler S. Longitudinal predictors of cannabis use and dependence in offspring from families at ultra high risk for alcohol dependence and in control families. Am J Med Genet Part B Neuropsychiatr Genet Off Publ Int Soc Psychiatr Genet. 2016;171B(3):383–395. doi:10.1002/ajmg.b.32417

77. Chorlian DB, Rangaswamy M, Manz N, et al. Gender modulates the development of theta event related oscillations in adolescents and young adults. Behav Brain Res. 2015;292:342–352. doi:10.1016/j.bbr.2015.06.020

78. Steffensen SC, Ohran AJ, Shipp DN, Hales K, Stobbs SH, Fleming DE. Gender-selective effects of the P300 and N400 components of the visual evoked potential. Vision Res. 2008;48(7):917–925. doi:10.1016/j.visres.2008.01.005

79. Koolschijn PCMP, Crone EA. Sex differences and structural brain maturation from childhood to early adulthood. Dev Cogn Neurosci. 2013;5:106–118. doi:10.1016/j.dcn.2013.02.003

80. Giedd JN, Blumenthal J, Jeffries NO, et al. Brain development during childhood and adolescence: a longitudinal MRI study. Nat Neurosci. 1999;2(10):861–863. doi:10.1038/13158

81. Troup LJ, Bastidas S, Nguyen MT, Andrzejewski JA, Bowers M, Nomi JS. An Event-Related Potential Study on the Effects of Cannabis on Emotion Processing. Felmingham K, ed. PLOS ONE. 2016;11(2):e0149764. doi:10.1371/journal.pone.0149764

82. D’Souza DC, Fridberg DJ, Skosnik PD, et al. Dose-Related Modulation of Event-Related Potentials to Novel and Target Stimuli by Intravenous Δ9-THC in Humans. Neuropsychopharmacology. 2012;37(7):1632–1646. doi:10.1038/npp.2012.8

83. Kempel P, Lampe K, Parnefjord R, Hennig J, Kunert HJ. Auditory-Evoked Potentials and Selective Attention: Different Ways of Information Processing in Cannabis Users and Controls. Neuropsychobiology. 2003;48(2):95–101. doi:10.1159/000072884

84. Solowij N, Michie PT, Fox AM. Effects of long-term cannabis use on selective attention: An event-related potential study. Pharmacol Biochem Behav. 1991;40(3):683–688. doi:10.1016/0091-3057(91)90382-C

85. Patrick G, Straumanis JJ, Struve FA, Fitz-Gerald MJ, Manno JE. Early and Middle Latency Evoked Potentials in Medically and Psychiatrically Normal Daily Marihuana Users: A Paucity of Significant Findings. Clin Electroencephalogr. 1997;28(1):26–31. doi:10.1177/155005949702800105

86. Hart CL, Ilan AB, Gevins A, et al. Neurophysiological and cognitive effects of smoked marijuana in frequent users. Pharmacol Biochem Behav. 2010;96(3):333–341. doi:10.1016/j.pbb.2010.06.003

