## Supplemental Materials for "Bidirectional associations between cannabis use, oddball performance and P3 event-related potential"

Sample description: The initial COGA sample consisted of individuals with alcohol use disorder (AUD) and their densely affected as well as unaffected family members as well comparison families with population-level liability to AUD. In 2004, the COGA Prospective cohort was recruited, comprising adolescent and young adult (ages 12-22 at their baseline assessment) offspring of individuals from earlier phases of COGA. These individuals were followed every 2 years with behavioral, clinical and neurocognitive assessments. A subsequent related component, the Lifespan Study, followed those aged 30 - 40 years from the Prospective Study and prior phases of COGA.

**eFigure 1.** Sex Stratified Post-onset P3 and Performance Trajectories by Cannabis Use Initiation

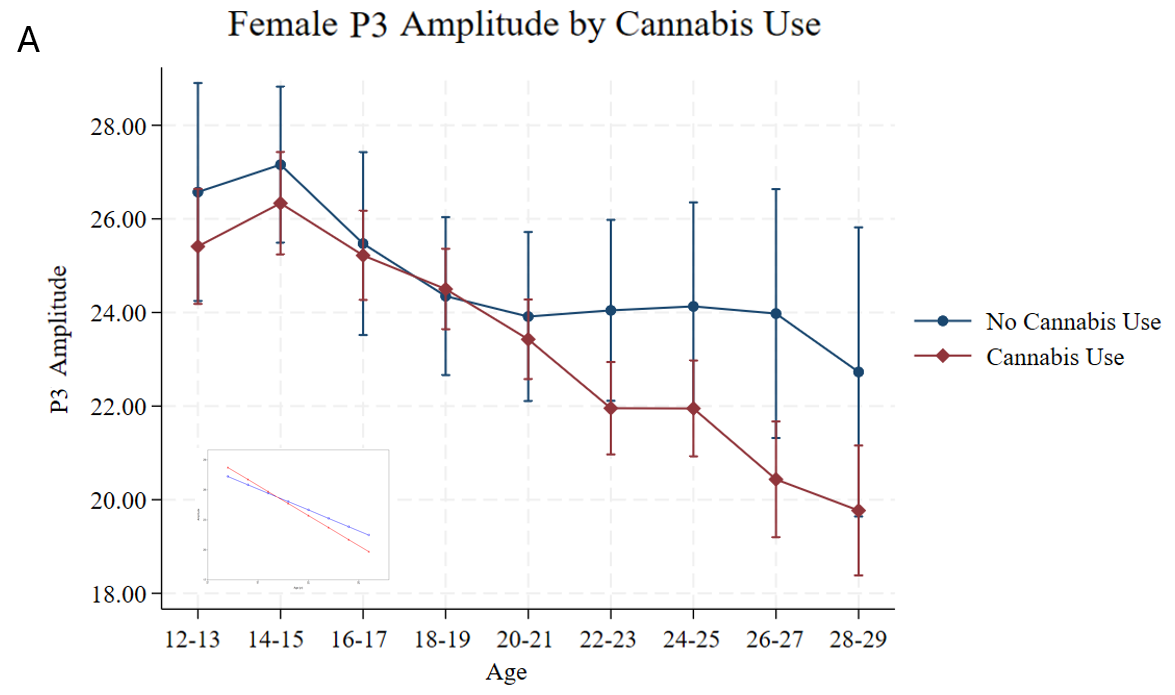

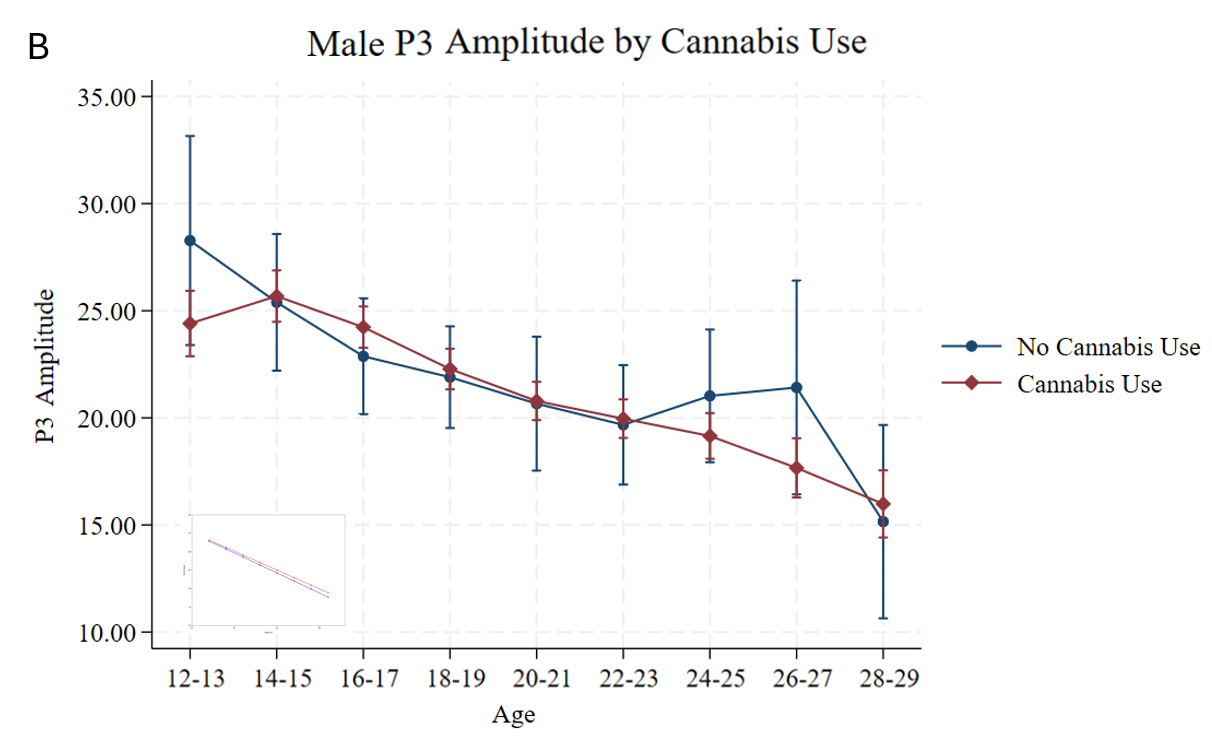

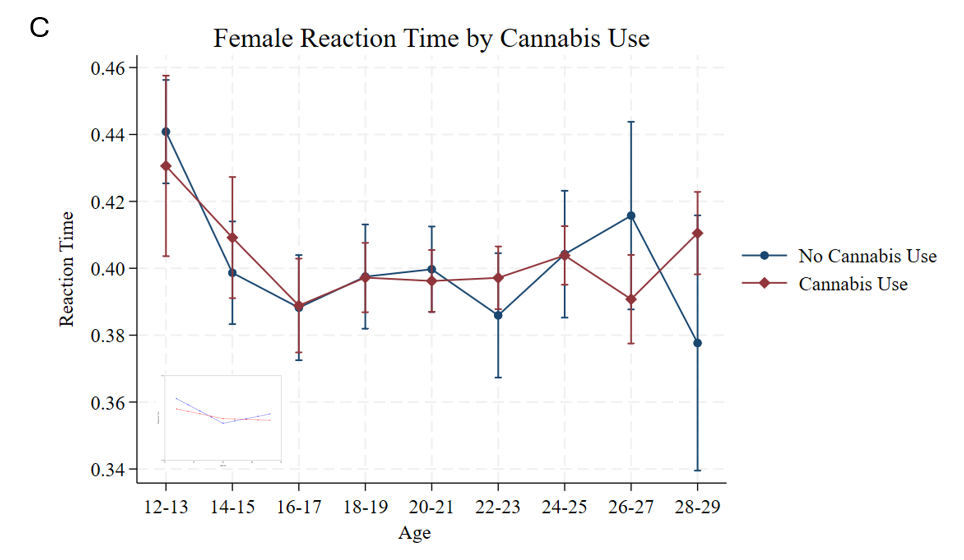

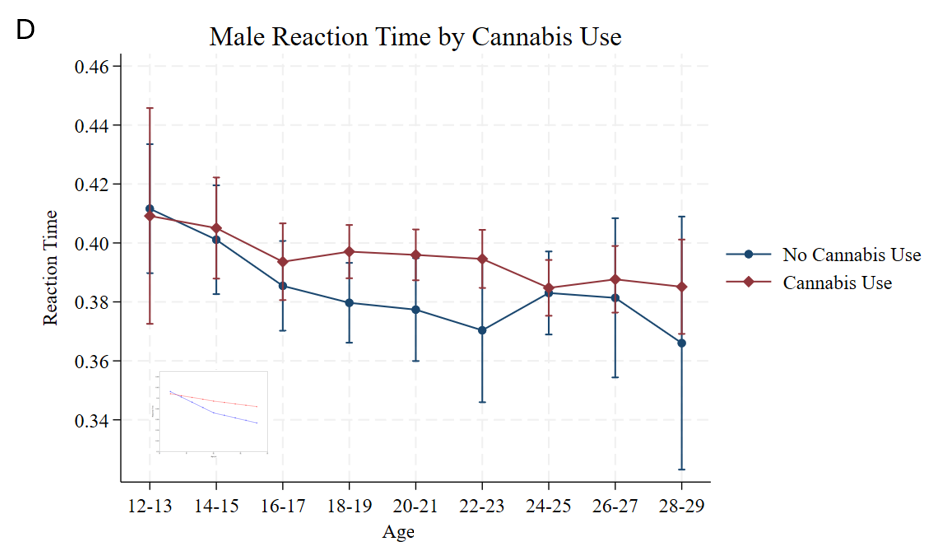

Sex stratified unadjusted mean changes in P3 amplitude (A & B) and reaction time (C & D) from ages 12 to 29 by cannabis use. Error bars represent 95% confidence intervals (CI). The inset panels display the selected latent growth curve adjusted for covariates

**eTable 1.** Time-varying Cannabis Models effect on P3 amplitude and Reaction Time

| **Age** | **B** | **CI** | **p** |
| --- | --- | --- | --- |
| Amplitude |  |  |  |
| 12-13 | -0.05 | -0.11, 0.01 | 0.162 |
| 14-15 | 0.04 | -0.01, 0.08 | 0.205 |
| 16-17 | 0.01 | -0.02, 0.05 | 0.522 |
| 18-19 | -0.03 | -0.06, 0.00 | 0.123 |
| **20-21** | **-0.04** | **-0.08, -0.01** | **0.025** |
| **22-23** | **-0.07** | **-0.10, -0.03** | **0.002** |
| **24-25** | **-0.06** | **-0.10, -0.02** | **0.013** |
| **26-27** | **-0.10** | **-0.15, -0.06** | **< .001** |
| **28-29** | **-0.12** | **-0.18, -0.07** | **.001** |
| Response Time |  |  |  |
| 12-13 | 0.08 | 0.04, 0.13 | 0.004 |
| 14-15 | -0.03 | -0.08, 0.01 | 0.228 |
| 16-17 | -0.05 | -0.09, -0.02 | 0.013 |
| 18-19 | -0.02 | -0.05, 0.01 | 0.246 |
| 20-21 | 0.01 | -0.02,0.04 | 0.604 |
| **22-23** | **0.04** | **0.00, 0.07** | **0.048** |
| **24-25** | **0.08** | **0.03, 0.12** | **0.005** |
| **26-27** | **0.07** | **0.02, 0.11** | **0.012** |
| **28-29** | **0.10** | **0.05, 0.15** | **0.002** |

Standardized estimates reflect the time varying association between cannabis use and P3 amplitude and reaction time at each 2-year age interval, with 95% confidence intervals (CI). Significant associations (p < .05) are shown in bold.
